# Context Matters in LLM-Assisted Qualitative Data Analysis: Workflow Development and Multidimensional Evaluation in Health Research

**DOI:** 10.64898/2026.09.07.26362410

**Authors:** Huohuo Dai, Yiling Li, María Villalobos, Yee Man Ng, Jiska J. Aardoom, Anke Versluis, Hongxia Shen, Marco Spruit, Niels H. Chavannes

## Abstract

Large language models (LLMs) are increasingly used for qualitative analysis, but strong language performance does not guarantee strong interpretation when meaning depends on method and context. We developed a context-specific workflow that translated framework analysis into bounded, sequential LLM-supported tasks and applied it to 20 Chinese-language interviews on fear of dementia (approximately 150,000 characters). Four multilingual LLM-generated outputs and a previous researcher-only analysis were blindly evaluated by 16 relevant experts across different analytical domains and qualitative quality dimensions (496 expert-item records). All LLMs completed the workflow, but rankings varied markedly. Qwen-output showed the most consistently favourable overall profile, while Claude-output was most often ranked first in individual domain–dimension evaluations. The researcher analysis received both highly favourable and highly unfavourable rankings; no output was consistently preferred. LLM-assisted qualitative analysis should be evaluated within its research contexts, with transparent workflows, multidimensional assessment, and continued researcher oversight.

## Introduction

Narrative data from interviews, focus groups, and open-ended responses are central to medical and health research, among many other disciplines. They show how people understand and experience illness, treatment and care in ways that are often difficult to capture through standardised measures [1–5]. Qualitative evidence is therefore widely used in patient-centred research, health-services research, intervention development, and implementation research[1,3]. However, analysing qualitative data requires sustained engagement with source material and interpretive judgement. Coding, comparing, and synthesising these data can consequently be time- and resource-intensive, particularly as datasets increase in size [3,6,7].

Large language models (LLMs) are creating new possibilities for supporting this work [8,9]. LLMs can process large volumes of natural-language text and may assist with tasks such as identifying relevant excerpts, generating candidate subthemes, and synthesising patterns across interviews [10–12]. Early studies have reported substantial reductions in processing time and have shown that LLMs can identify recurring structures in qualitative data [12–14]. At the same time, LLM-generated analyses can vary with model choice, prompt design, and contextual information, while long-context processing, reproducibility, and the interpretation of implicit or culturally situated meaning remain important concerns [15–18]. General language capability therefore offers no guarantee of high-quality qualitative interpretation [15]. Recent work has begun to incorporate richer methodological information into LLM-supported qualitative analysis [19]. However, less attention has been paid to designing the analytical workflow around the requirements and contexts of the qualitative study itself, such as its research question, analytical method, sociocultural setting, and intended purpose.

Evaluation presents a related challenge. Existing studies provide useful early evidence, but most evaluate LLM-assisted qualitative analysis using a relatively narrow set of outcomes, such as coding agreement, thematic overlap, or analytical speed [12–14,20–23], while emerging work has begun to consider broader dimensions of interpretive quality [19]. Qualitative analytical quality, however, cannot be captured by a single criterion or by correspondence with a pre-existing human analysis alone.

Methodological orientation and analytical purpose can partly be specified through the research question and analytical instructions, but qualitative interpretation also involves researcher expertise, contextual knowledge, reflexivity, and positionality [4,24]. Evaluation therefore needs to move beyond simple correspondence and consider the broader quality and defensibility of qualitative interpretation, including its grounding in the data, sensitivity to context, methodological coherence, and relevance to the intended research purpose [25]. To our knowledge, evidence comparing LLM-generated and researcher analyses across multiple dimensions of qualitative quality and across different analytical domains remains limited.

Framework analysis provides a particularly useful setting for such evaluation. It combines a structured and transparent analytical process with iterative interpretation, allowing deductive use of a conceptual framework to be integrated with inductive development of meanings grounded in participants’ accounts [26,27]. This makes the framework Analysis a useful methodological case for examining how an established qualitative method can be translated into explicit LLM-supported analytical tasks without reducing qualitative analysis to coding alone. However, most empirical studies of LLM-assisted qualitative analysis have focused on thematic analysis or coding [12–14,20,21], and we found little evidence examining their use within a framework analysis workflow. Research using non-English qualitative data is also much less common. Additionally, a further gap concerns culturally situated interpretation. For example, meanings related to family roles, social obligations, stigma, indirect expression, and illness beliefs may depend heavily on local sociocultural context, yet such contexts are less represented in current evaluations of LLM-assisted qualitative analysis [1,18,28–30].

Therefore, we aimed to develop and evaluate a context-specific LLM workflow for framework-informed qualitative analysis in health research. The workflow translates the steps of a framework analysis into bounded and sequential LLM-supported tasks. Context specificity referred to the deliberate alignment of the workflow with the research question, analytical method, the source language and relevant sociocultural context, the structure of the source data, and the intended analytical use of the findings. We operationalised framework analysis as a structured sequence of LLM-supported tasks and compared outputs from four LLM-generated outputs and a researcher-only analysis together through source-data verification and blinded multidimensional expert evaluation. We used Chinese-language interviews on fear of dementia as an information-rich methodological case. The interviews contained emotionally complex and culturally situated accounts of autonomy, stigma, family responsibility, coping, and future care, and the study questions were well suited to the structured yet interpretive approach of the framework analysis method [29,31–34].

## Methods

### Study design

We conducted a comparative methodological study to develop and evaluate a context-specific, LLM-assisted workflow for framework analysis of Chinese-language health interviews. The study comprised two linked components: (1) iterative development of a context-specific LLM-assisted analytical workflow; and (2) blinded expert evaluation of a pre-existing researcher-only output and four LLM-generated outputs produced using the LLM-workflow. Before expert evaluation, the four LLM-generated outputs underwent researcher verification of quotation accuracy, participant attribution, domain relevance, and evidentiary support.

The comparative evaluation included five final analytical outputs derived from the same interview dataset and informed by the same conceptual framework: one pre-existing researcher-only analysis and four outputs generated by different multilingual LLMs using the same LLM-assisted workflow. The researcher-only analysis had been completed independently before any LLM-generated outputs were produced and was not treated as a definitive gold standard. The LLM-assisted workflow was finalised before formal model execution and applied without model-specific optimisation. The study reports analytical performance observed under these predefined workflow conditions.

### Data source and preparation

We reused a collection of 20 anonymous, Chinese-language verbatim transcripts from semi-structured interviews with middle-aged and older adults in China. The dataset comprised 148,195 Chinese characters. The transcripts had been proofread and checked by the research team for transcription accuracy and coherence before being reused in the present methodological study. To preserve comparability with the pre-existing researcher-only analysis, transcripts were prepared according to a minimal-transformation principle. All transcripts were anonymized before being uploaded to the LLM platforms. Names, contact details, institutional identifiers, specific locations, and other potentially identifying personal information were removed or generalised. No textual content, question order, paragraph structure, or participant responses were otherwise edited, summarised, or reorganised. The original MS Word files were converted to UTF-8-encoded text files, with one transcript per participant and anonymous study identifiers retained solely to support evidence tracing. Further details on the data source and transcript preparation are provided in the ***Supplementary Methods***.

### Pre-existing researcher-only analysis

Full details of the researcher-only analysis and researcher reflexivity are reported in previously [35] and ***Supplementary Materials***. Briefly, two researchers analysed the transcripts using the framework analysis, informed by the Theoretical Domains Framework (TDF) [27,36]. After familiarisation and pilot coding, they applied the TDF deductively while developing inductive subthemes grounded in participants’ accounts. Coding decisions and emerging interpretations were refined through iterative discussion within the multidisciplinary research team, supported by analytic notes and an audit trail. The researcher-only analysis was included as one of the five blinded analytical outputs for comparative expert evaluation. The five outputs were evaluated blindly using the same prespecified criteria.

### Design of the context-specific LLM-assisted workflow

We developed a structured, practical context-specific workflow to translate the main stages of Framework Analysis into bounded tasks suitable for LLM processing. The workflow was informed by the analytical approach and requirements of the parent study [35]together with practical constraints associated with long-context LLM use.

The workflow was iteratively developed by two researchers (HD and YL). The initial version used a single comprehensive prompt that combined contextual information with multiple analytical tasks. Although the models were able to generate complete outputs, researcher verification during pilot testing identified limitations in analytical quality and consistency, including uneven coverage of the source data and supporting evidence drawn disproportionately from earlier parts of the input. These observations suggested that contextual specification alone was insufficient; the analytical work also needed to be divided into more manageable tasks [18,30]. We therefore decomposed the analysis into sequential prompts and revised the input strategy to improve evidence coverage and consistency during long-context processing.

The final workflow comprised two set-up prompts and four analytical prompts. The set-up prompts first established the model’s analytical role and provided the research context (e.g., background, setting and culture, research questions, interview guide, TDF definitions, output requirements, and evidence-traceability rules). The analytical prompts then guided the model through four steps: identifying excerpts relevant to the research questions, assigning these excerpts deductively to TDF domains with supporting rationale, developing data-grounded subthemes within each domain, and synthesising the resulting subthemes across transcript groups while retaining participant-level evidence and important differences or contradictions. This structure was intended to keep LLM involvement tied to specific analytical tasks. The analytical workflow is shown in ***Fig. 1***, and the complete prompts and detailed procedures are provided in the ***supplementary materials***.

**Figure 1.**
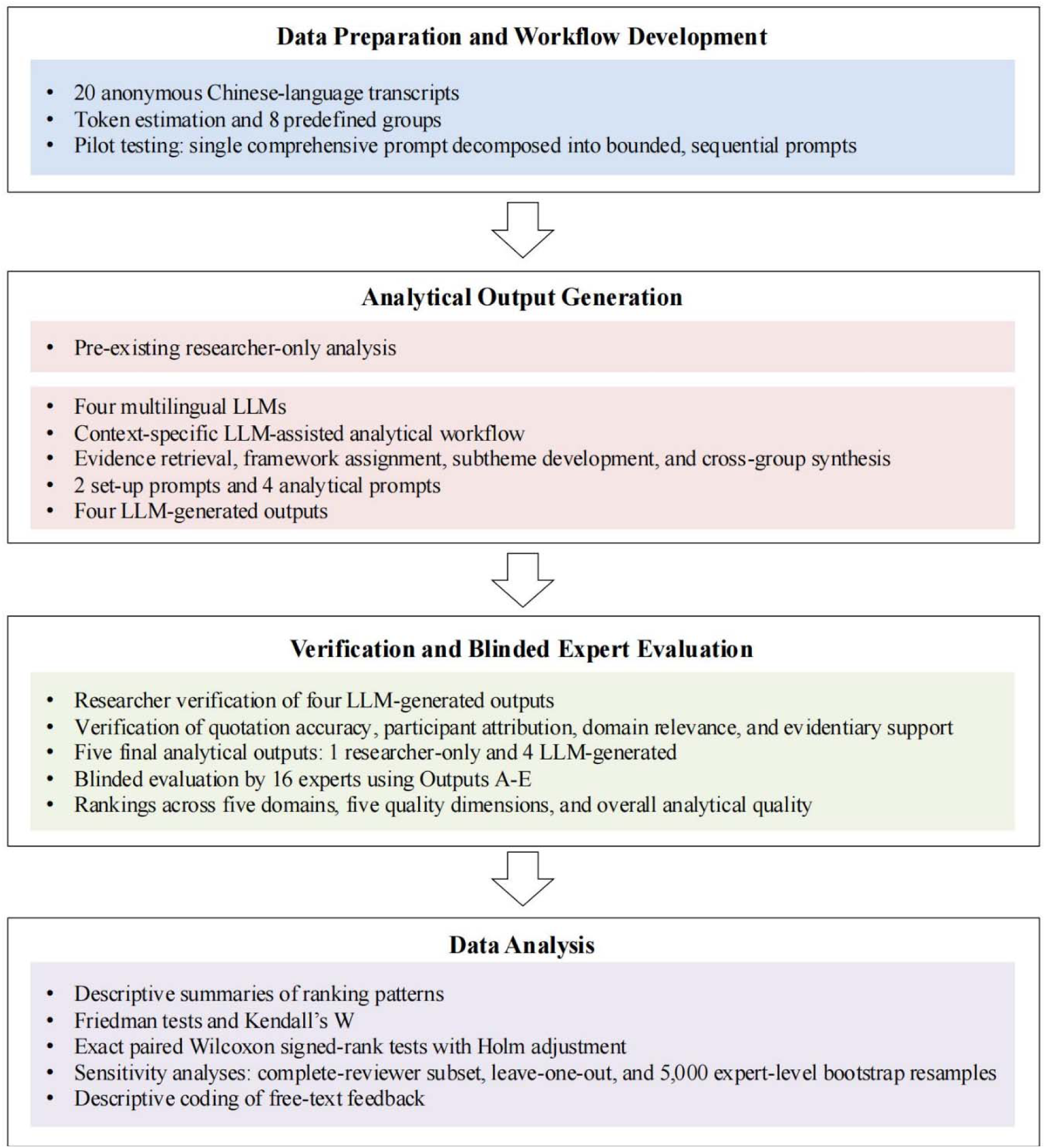
Overview of the study procedure

### Workflow Standardisation and Input Management

Transcript length was estimated using the cl100k_base tokenizer as a common approximation for input-capacity planning. The 20 transcripts were allocated to eight predefined groups, with individual transcripts kept intact and group composition and input order held constant across all four models. Most groups contained approximately 20,000–22,000 estimated tokens. This range was used as a pragmatic planning threshold during workflow development to limit long-context input while maintaining consistent grouping across models; it was not intended as a model-specific context-window limit. Full grouping details are provided in the ***Supplementary Materials***.

Each transcript group was analysed in a separate conversation using the same set-up and analytical prompts. Group-level outputs were subsequently used for cross-group synthesis. Following pilot testing, the prompt wording and sequence, transcript grouping, input procedures, output templates, and permissible corrective instructions were fixed before formal execution. All prompts were administered in Chinese, and no model-specific prompt optimisation was undertaken. If an output was interrupted because of platform or length constraints, the only permitted instruction was to continue the original task without changing its analytical requirements.

### LLM selection

We selected four general-purpose multilingual LLMs—ChatGPT (GPT-5.5), Claude (Claude Opus 4.8), DeepSeek (DeepSeek V4), and Qwen (Qwen3.7)—because they supported Chinese-language input, were publicly accessible during the study period, and represented widely used international and Chinese-developed model families. This selection reflected a pragmatic use case in which health researchers could conduct LLM-assisted analysis without specialist programming or local deployment infrastructure [37]. For each model, we documented the displayed model or platform label, access date, interface, subscription mode, relevant privacy settings, and operational constraints.

### Researcher verification of output fidelity

Before blinded expert evaluation, the LLM-generated outputs underwent researcher verification of features that could be checked directly against the source data. The original interview transcripts served as the primary reference for verifying quotation authenticity, participant identifiers, and evidentiary support. Verification examined whether extracted excerpts were reproduced accurately, attributed to the correct participant, and relevant to the assigned framework domain, and whether generated subthemes and cross-case claims were supported by the cited data. This verification also allowed us to identify potential hallucination-related problems, such as fabricated, misattributed, or unsupported evidence. Researcher verification was used to characterise output fidelity and recurring failure modes; it was not used to correct the outputs subsequently presented to experts.

### Blinded expert evaluation

We conducted a blinded comparative evaluation of the five final analytical outputs: one pre-existing researcher-only analysis and four LLM-generated outputs. Each output represented the final cross-group synthesis and included domain-level subthemes, their definitions or core meanings, and supporting participant-level evidence. Experts were purposively recruited to provide complementary expertise in qualitative research, data science, implementation or behavioural science, AI-assisted research, and culturally informed Chinese health research.

Experts assessed outputs from five prespecified analytical framework domains: Emotion, Beliefs about Consequences, Social Influences, Environmental Context and Resources, and Knowledge. Within each domain, the five outputs were presented in a standardised format and labelled only as Outputs A-E, with model or researcher identities concealed. Experts ranked the outputs comparatively rather than scoring each one independently. Within each domain, experts first ranked the five outputs from strongest to weakest according to overall analytical quality and then ranked them separately across five dimensions of qualitative analytical quality: interpretive depth, cultural and contextual sensitivity, evidence logic and traceability, framework fit, and practical usefulness for intervention design. Experts also provided an overall ranking of the five outputs; the output ranked first was considered the one most representative of high-quality framework-informed qualitative analysis. Optional free-text comments were also collected. A comparative ranking design was chosen to encourage discrimination between outputs and to reduce ceiling effects and limited variability associated with absolute rating scales. Further details and a representative example of the blinded evaluation materials are provided in the ***Supplementary Materials***.

### Data analysis

Quantitative analyses and visualisation were conducted using R (version 4.5.1) and Python (version 3.12). Operational records and expert characteristics were summarised descriptively. Researcher-verification findings were synthesised according to the type of output fidelity and integrated narratively.

Expert rankings were summarised using mean rank, median, interquartile range, rank distributions, and the frequencies with which each output was ranked first, within the top two, or last. Overall differences among outputs were examined using Friedman tests, and Kendall’s coefficient of concordance was used to assess agreement among experts. Exact paired Wilcoxon signed-rank tests with Holm adjustment were used for exploratory pairwise comparisons. Analyses examined overall, domain-specific, and quality-dimension-specific patterns. For each quality dimension, rankings were averaged across the five domains within each expert, and the resulting expert-level mean ranks were compared across outputs. First-place frequencies across the 80 repeated expert-domain evaluations were also summarised descriptively. Selection of the single output considered most representative of high-quality framework-informed qualitative analysis and predefined output limitations were summarised using frequencies and percentages.

Supplementary analyses included Sensitivity analyses and descriptive coding of free-text feedback. Sensitivity analyses included restrictions to the 10 experts who reported fully reviewing the supporting materials, leave-one-out analyses, and expert-level bootstrap resampling with 5,000 iterations. Free-text comments were reviewed and descriptively coded by two researchers (HD, YL) to identify recurring evaluative patterns. Given the limited volume of comments, coding was used to support interpretation.

### Ethical considerations and data governance

The parent interview study, from which the interview dataset was derived, was approved by the relevant Chinese ethics review committee (Guangzhou Medical University, approval number: 202504010). All participant data were collected, stored, and used in accordance with the ethical requirements of the parent study. The present study received separate ethics approval from Guangzhou Medical University (approval number: 202607008). Only anonymous transcripts were submitted to the LLM platforms. All direct identifiers and potentially identifying contextual details had been removed or generalised before model processing, and no personal information was uploaded. Data minimisation was applied so that models received only the transcript content and contextual information necessary to complete the predefined analytical tasks. The four models were accessed through publicly available cloud-based interfaces. LLM-generated outputs were stored within the research environment and used solely for methodological comparison. Platform-specific access and privacy conditions are summarised in the ***Supplementary Materials***.

## Discussion

This study developed and evaluated a context-specific LLM workflow for qualitative framework analysis and compared four LLM-generated analyses with a pre-existing researcher analysis using blinded, multidimensional expert evaluation. The workflow showed that an established qualitative method could be translated into a sequence of bounded LLM-supported tasks while retaining links to the study purpose, conceptual framework and source data. At the same time, the evaluation did not reveal a stable hierarchy among the five analyses. Relative performance changed across analytical domains and dimensions of qualitative analytical quality, and no output was consistently preferred. These findings suggest that the value of an LLM in qualitative research depends on how it is used within a particular analytical process and on how the resulting interpretation is judged.

The workflow was developed from the requirements of the qualitative study itself. The research context (e.g., research question, analytical method, source language, relevant sociocultural context, and intended analytical outputs) were specified before being translated into bounded analytical tasks. Task decomposition therefore served not only to manage model input and output, but also to preserve the analytical logic of the study and make intermediate analytical steps more transparent and traceable.

Emerging methodological work similarly argues for task-specific and researcher-directed uses of LLMs [19,38,39]. Our study builds on this direction by placing qualitative methodological requirements at the centre of workflow design and by evaluating the resulting outputs across dimensions that capture both evidentiary quality and interpretive depth. Applying the same workflow across four multilingual LLMs using original-language data further allowed these outputs to be examined within a common analytical structure. This distinction is increasingly relevant as generative AI functions become embedded within qualitative analysis software, where technical integration does not by itself ensure alignment with the methodological logic and interpretive requirements of a qualitative study [15].

The multidimensional evaluation showed that the relative strengths of the analytical outputs depended on what aspect of qualitative quality was being considered. Qwen showed the most consistent overall profile, whereas Claude received more first-place evaluations on individual quality assessments, particularly for interpretive depth and evidence logic and traceability. Expert comments reflected similar trade-offs between comprehensiveness and focus, granularity and fragmentation, interpretive depth and over-reading, and evidence density and thematic coherence. Previous comparative studies have shown that human and LLM analyses may identify overlapping findings while emphasising different aspects of the same qualitative data [12]. Other studies have reported substantial human–LLM agreement and efficiency gains [13,14,40]; recent work has also shown the value of assessing LLM-generated qualitative analyses against broader criteria of interpretive quality [19]. Our findings add a further layer by showing that relative quality was not stable across analytical domains or evaluative dimensions. This suggests that evaluating LLM-supported qualitative analysis only by its similarity to a human analysis, or by a single overall performance measure, may overlook important differences in interpretive quality.

The polarised evaluation of the pre-existing researcher analysis is also methodologically important. Although this analysis had been completed independently of the LLM outputs, it received both highly favourable and highly unfavourable rankings in the blinded evaluation. This finding cautions against treating a single human analysis as an uncontested reference standard. Qualitative interpretation is shaped by the analytical purpose, theoretical orientation, researcher expertise, contextual knowledge, and judgements about what is most salient in the data [3,4,24,25]. Some of these methodological elements can be made explicit within an LLM workflow, but researcher positionality and situated interpretive experience are not readily reducible to instructions [15]. Differences between the researcher analysis and an LLM-generated output therefore do not necessarily indicate analytical error on either side. They may also reflect legitimate differences in emphasis and interpretation. For comparative evaluation, this supports judging outputs against explicit criteria such as grounding in the source data, contextual and cultural sensitivity, evidentiary coherence, and methodological fit, rather than relying only on correspondence with a single human analysis.

Performance also varied across the evaluated framework structure. Expert rankings were more consistent for the Knowledge domain than for the Emotion domain. This may partly reflect differences in the material being interpreted: knowledge-related statements were often more explicit, whereas emotional accounts could depend more on nuance, relational context, and implicit meaning. These findings suggest that the difficulty of LLM-supported qualitative interpretation may vary across different types of content, even within the same study. This task-dependent view is consistent with broader medical AI literature, which increasingly emphasises matching the degree of AI involvement and human oversight to the characteristics and consequences of the task [41].

The use of original Chinese-language interviews for LLM analysis also highlights the need to distinguish linguistic capability from culturally situated interpretation. Analysing data in the source language may help preserve meanings that could otherwise be altered through translation, but multilingual fluency does not in itself ensure sensitivity to the social and cultural contexts in which those meanings are produced. In this study, narratives of fear of dementia were often embedded in family responsibility, relational expectations, stigma, and concerns about future dependence, all of which can shape how participants express and interpret their experiences [29,31]. Cultural and contextual sensitivity therefore represents more than accurate language processing. It requires attention to how meanings are situated within relationships, social roles, and local understandings of health and illness. For LLM-assisted qualitative analysis, language and sociocultural context should therefore be treated as related but distinct components of workflow design [28,38].

These findings make model-level rankings a limited basis for judging qualitative analytical value. A more useful question is whether a particular model–workflow configuration produces interpretations that are credible for the study in which it is used. The level of researcher involvement may therefore need to vary across tasks, especially as interpretation becomes less directly verifiable and more dependent on contextual judgement.

This has practical implications for how qualitative AI studies are designed and reported. Reporting should go beyond the model name and prompt. At minimum, studies should describe where the LLM entered the analytical process, what methodological and contextual information it received, how its outputs were checked, and which interpretive decisions remained with researchers [42,43]. As generative AI becomes increasingly embedded within qualitative analysis software, documenting these elements will help distinguish methodological integration from the simple availability of an AI function [10]. Such transparency also helps to clarify accountability: AI may contribute to the analytical process, but responsibility for how participants’ accounts are interpreted, represented, and translated into research claims remains with the research team. This is especially important in qualitative health research, where analytical interpretations can shape how participants’ experiences are represented and which research or intervention priorities receive attention.

A strength of this study is that all four LLMs were evaluated within the same predefined analytical workflow rather than through model-specific prompting. The blinded evaluation also moved beyond coding agreement or thematic overlap by comparing analyses across both analytical domains and dimensions of qualitative quality. Including a pre-existing researcher analysis within the same blinded comparison further allowed human interpretation to be examined rather than assumed to be the reference standard. The findings should be interpreted within the scope of this methodological case study. We evaluated one Chinese-language interview dataset using Framework Analysis, and the observed patterns may differ in other research contexts. The results also reflect the model versions and public interfaces available during the study period and should therefore be understood as evidence about the evaluated model–workflow configurations rather than fixed properties of particular model families. Finally, expert rankings captured relative judgements among the five analytical outputs and were not intended to establish an absolute threshold of qualitative quality.

In summary, this study developed and evaluated a context-specific LLM workflow for qualitative framework analysis and showed that the quality of resulting analytical outputs cannot be inferred from model identity alone. Across four LLM-generated outputs and a pre-existing researcher analysis, relative performance varied across analytical domains and dimensions of qualitative quality, with no single output consistently preferred. These findings support evaluating LLM-assisted qualitative analysis through the quality and defensibility of the interpretation it produces within a defined methodological and contextual setting. As LLMs become more deeply integrated into qualitative health research, technical capability should be matched by research contexts, methodological transparency and clear interpretive accountability, particularly when AI-generated interpretations contribute to the representation of patient experiences and subsequent scientific claims.

## Results

### Workflow execution and output verification

The iterative development process resulted in a six-prompt workflow comprising two set-up prompts and four sequential analytical prompts, with task decomposition and input management refined during pilot testing to improve evidence coverage and output consistency. Researcher verification indicated that the formal LLM outputs were generally traceable to the source transcripts, with no prominent pattern of fabricated quotations or participant misattribution; remaining concerns related mainly to the strength and appropriateness of analytical interpretation.

### Overall and domain-specific expert evaluation

Sixteen experts completed all 31 ranking items (six rankings within each of the five domains and one overall ranking across the five outputs), yielding 496 valid expert-item records with no missing or duplicated ranks. Expert characteristics and a representative example of the domain-level evaluation are provided in the ***Supplementary Materials***. Overall rankings (***Table 1***) differed across the five outputs (Friedman χ² (4) =16.60, p=.002), with limited expert concordance (Kendall’s W=.259; ***Table 1***). Qwen showed the most favourable overall ranking profile. The researcher-only analysis showed the most dispersed profile. After Holm adjustment, only the Qwen-ChatGPT comparison remained statistically significant (adjusted p=.007).

**Table 1.** Overall expert ranking profiles of the five analytical outputs.

| Analytical output | Mean rank (SD) | Median (IQR) | Ranked first, n (%) | Top two, n (%) | Ranked fifth, n (%) |
| --- | --- | --- | --- | --- | --- |
| ChatGPT | 4.12 (0.89) | 4.00 (4.00-5.00) | 0 (0.0) | 1 (6.3) | 6 (37.5) |
| **Qwen** | **2.00 (1.10)** | **2.00 (1.00-2.00)** | **6 (37.5)** | **13 (81.3)** | **0 (0.0)** |
| DeepSeek | 3.12 (1.02) | 3.00 (3.00-3.25) | 1 (6.3) | 3 (18.8) | 2 (12.5) |
| Researcher-only analysis | 3.25 (1.61) | 3.50 (1.75-5.00) | 4 (25.0) | 5 (31.3) | 5 (31.3) |
| Claude | 2.50 (1.51) | **2.00 (1.00-3.25)** | 5 (31.3) | 10 (62.5) | 3 (18.8) |

*Note: Ranks ranged from 1 (most favourable) to 5 (least favourable). Friedman chi-square (4)=16.60, $p$ =0.002; Kendall's $W$ =0.259. After Holm adjustment, only Qwen versus ChatGPT remained significant (adjusted $p$ =0.007). Bold values indicate the most favourable value for each ranking indicator; ties are both shown in bold.*

When domain-specific rankings were averaged across the five domains within each expert, significant differences remained among the five outputs, Friedman χ² (4)=21.37, p<.001, with moderate expert concordance (Kendall’s W=.334; ***Table 2***). The Qwen-generated output showed the most favourable across-domain ranking profile. After Holm adjustment, Qwen (adjusted p=.002) and DeepSeek (adjusted *p*<.001) were ranked more favourably than ChatGPT. No other pairwise comparison remained statistically significant. Domain-specific patterns varied (***Table 2*, *Figure 2***). The Qwen-generated output had the most favourable mean rank in Emotion, Social influences, and Environmental context and resources. The DeepSeek-generated output ranked most favourably in Beliefs about consequences, while the Qwen- and Claude-generated outputs were tied in Knowledge. Expert concordance also varied across domains (***Table 2***). Expert evaluations were therefore more consistent for Knowledge than for Emotion.

**Figure 2.**
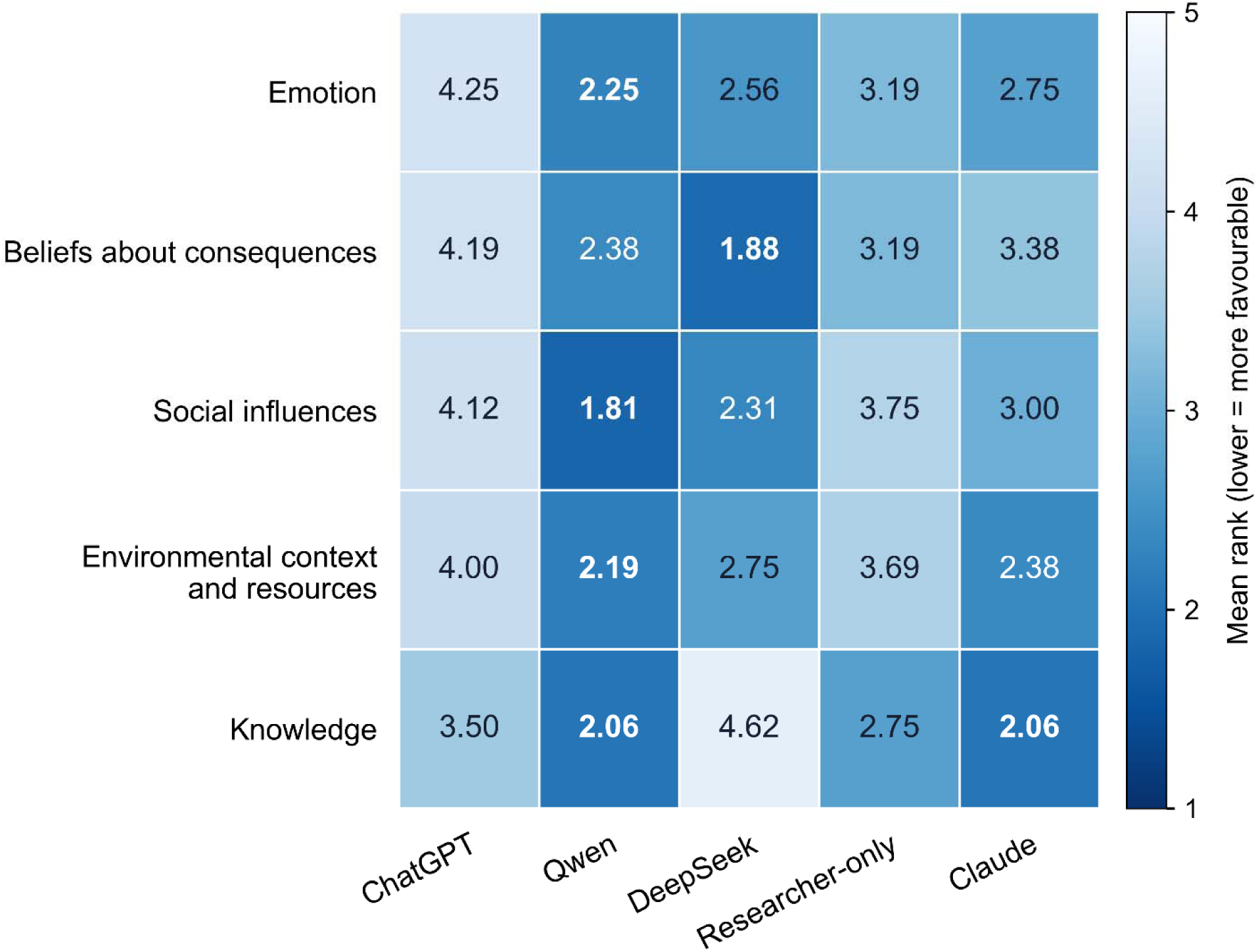
Domain-specific mean rankings of the five analytical outputs

**Table 2.** Domain-specific expert rankings of the five analytical outputs.

| Domain | Chat GPT | Qwen | DeepSeek | Researcher-only | Claude | Friedman chi-square | p | Kendall's W |
| --- | --- | --- | --- | --- | --- | --- | --- | --- |
| Emotion | 4.25 (1.13) | **2.25 (0.93)** | 2.56 (1.41) | 3.19 (1.33) | 2.75 (1.48) | 15.45 | 0.004 | 0.241 |
| Beliefs about consequences | 4.19 (1.11) | 2.38 (1.15) | **1.88 (0.81)** | 3.19 (1.38) | 3.38 (1.45) | 20.75 | <0.001 | 0.324 |
| Social influences | 4.12 (0.89) | **1.81 (1.05)** | 2.31 (1.01) | 3.75 (1.29) | 3.00 (1.46) | 23.75 | <0.001 | 0.371 |
| Environmental context and resources | 4.00 (1.21) | **2.19 (1.42)** | 2.75 (0.93) | 3.69 (1.25) | 2.38 (1.41) | 16.55 | 0.002 | 0.259 |
| Knowledge | 3.50 (1.03) | **2.06 (0.85)** | 4.62 (0.89) | 2.75 (1.24) | **2.06 (1.24)** | 30.15 | <0.001 | 0.471 |
| **Across five domains** | 4.01 (0.67) | **2.14 (0.85)** | 2.83 (0.53) | 3.31 (1.15) | 2.71 (1.21) | 21.37 | <0.001 | 0.334 |

*Note: Values are mean rank (SD); lower values indicate more favourable evaluations. The final row summarizes expert-level mean ranks across the five domains. In the across-domain analysis, Qwen and DeepSeek were ranked more favourably than ChatGPT after Holm adjustment (adjusted $p=0.002$ and $p<0.001$ , respectively). Bold values indicate the most favourable mean rank within each domain; ties are both shown in bold.*

### Multidimensional evaluation of qualitative quality

The five analytical outputs also differed across all five dimensions of analytical quality (***Table 3;* *Figure 3***). The Qwen-generated output had the most favourable mean-ranking profile for interpretive depth, cultural and contextual sensitivity, framework fit, and practical usefulness for intervention design, whereas the Claude-generated output ranked most favourably for evidence logic and traceability.

**Figure 3.**
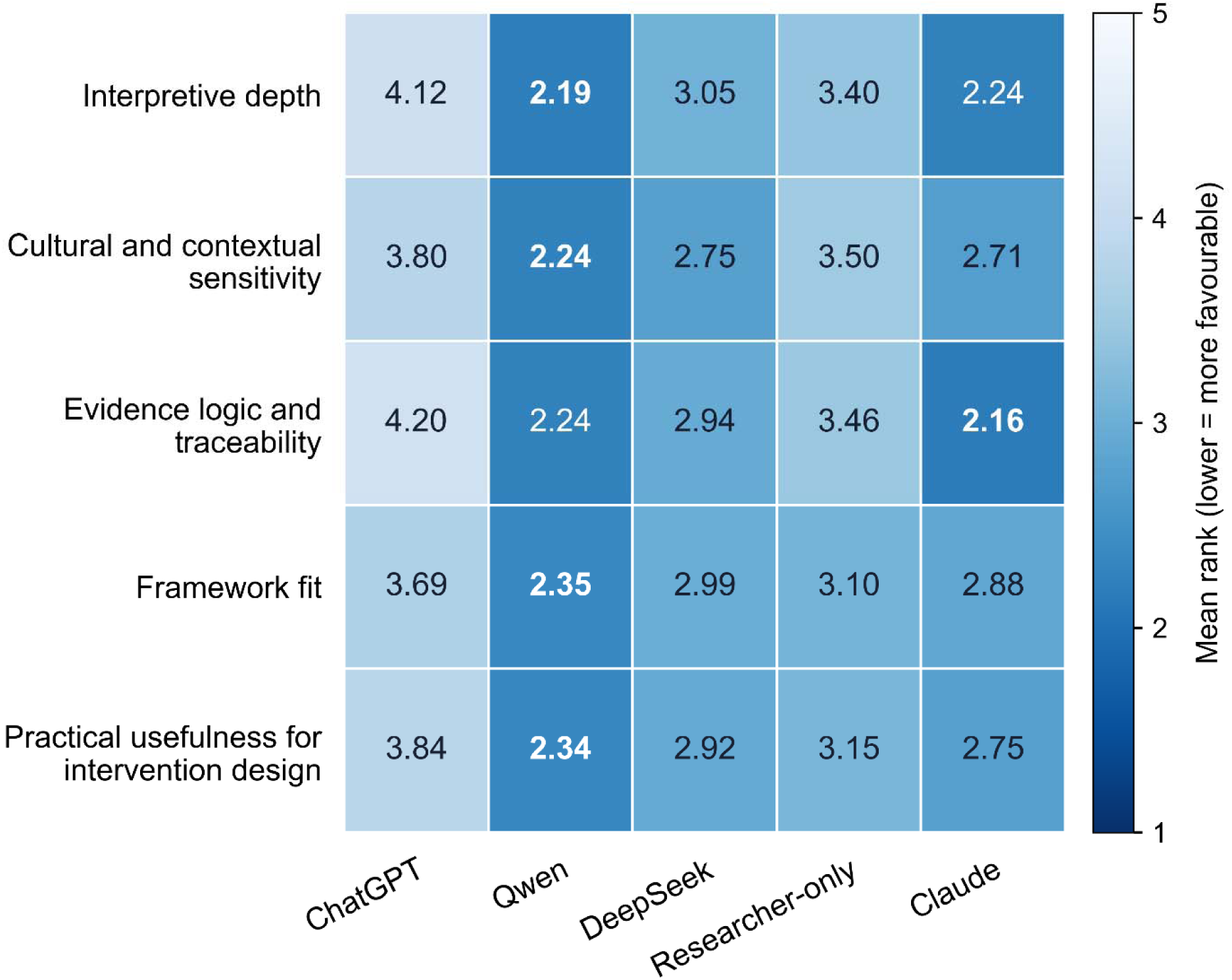
Multidimensional quality ranking of the five analytical outputs

**Table 3.** Expert rankings of the five analytical outputs across five dimensions of qualitative analytical quality.

*Panel A. Expert mean ranks and inferential statistics*
| Quality dimension | ChatGPT | Qwen | DeepSeek | Researcher-only | Claude | Friedman chi-square | p | Kendall's W |
| --- | --- | --- | --- | --- | --- | --- | --- | --- |
| Interpretive depth | 4.12 | **2.19** | 3.05 | 3.40 | 2.24 | 25.87 | <0.001 | 0.404 |
| Cultural and contextual sensitivity | 3.80 | **2.24** | 2.75 | 3.50 | 2.71 | 20.55 | <0.001 | 0.321 |
| Evidence logic and traceability | 4.20 | 2.24 | 2.94 | 3.46 | **2.16** | 35.11 | <0.001 | 0.549 |
| Framework fit | 3.69 | **2.35** | 2.99 | 3.10 | 2.88 | 15.85 | 0.003 | 0.248 |
| Practical usefulness for intervention design | 3.84 | **2.34** | 2.92 | 3.15 | 2.75 | 12.54 | 0.014 | 0.196 |

*Panel B. Descriptive first-place distributions across repeated expert-domain evaluations*
| Quality dimension | ChatGPT, n/80 (%) | Qwen, n/80 (%) | DeepSeek, n/80 (%) | Researcher-only, n/80 (%) | Claude, n/80 (%) |
| --- | --- | --- | --- | --- | --- |
| Interpretive depth | 2/80 (2.5) | 20/80 (25.0) | 9/80 (11.3) | 12/80 (15.0) | **37/80 (46.3)** |
| Cultural and contextual sensitivity | 5/80 (6.3) | 23/80 (28.8) | 18/80 (22.5) | 7/80 (8.8) | **27/80 (33.8)** |
| Evidence logic and traceability | 1/80 (1.3) | 21/80 (26.2) | 13/80 (16.3) | 6/80 (7.5) | **39/80 (48.8)** |
| Framework fit | 4/80 (5.0) | 21/80 (26.3) | 11/80 (13.8) | 20/80 (25.0) | **24/80 (30.0)** |
| Practical usefulness for intervention design | 4/80 (5.0) | 21/80 (26.3) | 14/80 (17.5) | 16/80 (20.0) | **25/80 (31.3)** |

Note: Panel A uses one expert-level observation per expert (n=16), calculated by averaging the five domain-specific rankings within each expert and output. Panel B is descriptive because the 80 records are repeated evaluations from 16 experts rather than 80 independent observations. Holm-adjusted pairwise comparisons across all output pairs are reported in the Supplementary Materials. In Panel A, bold values indicate the most favourable mean rank within each quality dimension. In Panel B, bold values indicate the highest first-place frequency within each quality dimension.

Across the repeated expert-domain evaluations, the Claude-generated output received the largest number of first-place rankings in each of the five quality dimensions, ranging from 24 of 80 evaluations (30.0%) to 39 of 80 (48.8%; ***Table 3***). These distributions were interpreted descriptively because the 80 evaluations represented repeated judgements from 16 experts. Overall, the Qwen-generated output showed a more consistently favourable mean-ranking profile, whereas the Claude-generated output was more frequently ranked first in individual domain-dimension evaluations.

In the overall ranking of the five outputs, experts most often selected Claude-generated output as the best represented high-quality qualitative analysis (5/16, 31.3%), followed by Qwen-generated output and the researcher-only analysis (4/16, 25.0% each), and DeepSeek-generated output (3/16, 18.8%). ChatGPT-generated output was not selected once. No output achieved majority support, indicating no clear consensus on a single preferred analytical output.

### Sensitivity analyses

Sensitivity analyses yielded patterns consistent with the primary findings. Free-text comments identified recurring evaluative patterns concerning comprehensiveness, granularity, interpretive depth, and evidentiary coherence. Detailed supplementary analyses are reported in the ***Supplementary Materials***.

## Declarations

### Data availability

The interview transcripts generated and analysed during this study are not publicly available because they contain potentially identifiable and sensitive qualitative information and their public sharing was not covered by participant consent. De-identified data supporting the findings may be available from the corresponding author upon reasonable request, subject to ethical and institutional requirements.

### AI usage disclosure statement

Large language models (LLMs) were used as part of the study workflow, as described in the Methods and Supplementary Materials. During manuscript preparation, ChatGPT 5.5 was used only to assist with language refinement, clarity, and editing of author-written text. All AI-assisted text was critically reviewed and revised by the authors, who take full responsibility for the accuracy, integrity, and final content of the manuscript.

### Author contributions

Conceptualization: HD, NC; Data curation: HD, YL; Formal analysis: HD, YL; Funding acquisition: HD, NC, MS; Investigation: HD, YL; Methodology: HD, YL, NC, MS; Project administration: HD; Resources: HD, NC; Supervision: NC, MS; Validation: HD, YL; Visualization: YL; Writing – original draft: HD, YL; Writing – review & editing: all authors

## Acknowledgments

The authors thank Xin Zhang (School of Biomedical Engineering & Imaging Sciences, King’s College London), Yu Wang (Department of Pain Medicine, Zhongnan Hospital of Wuhan University), Jiajia Liu and Wenjun Chen (Xiangya School of Nursing, Central South University), Beilei Lin (School of Nursing and Health, Zhengzhou University), Gang Gan, and ten other anonymous experts for their valuable contributions to the expert evaluation conducted in this study.

